# The Positive F Wave in Lead V1 of Typical Atrial Flutter is Caused by Activation of the Right Atrial Appendage: Insight from Mapping During Entrainment from the Right Atrial Appendage

**DOI:** 10.1101/2023.10.09.23296779

**Authors:** Shu Yamashita, Akira Mizukami, Maki Ono, Jiro Hiroki, Shota Miyakuni, Hina Miyamoto, Hiroki Ishizu, Takumi Arashiro, Soichi Noguchi, Daisuke Ueshima, Akihiko Matsumura, Shinsuke Miyazaki, Tetsuo Sasano

**Affiliations:** Department of Cardiology, National Hospital Organization Disaster Medical Center, Tokyo, Japan; Department of Cardiology, Kameda Medical Center, Chiba, Japan; Department of Medical Engineering, Kameda Medical Center, Chiba, Japan; Department of Cardiovascular Medicine, Tokyo Medical and Dental University, Tokyo, Japan

**Author notes:** Address for correspondence: Akira Mizukami, MD., Department of Cardiology, Kameda Medical Center, Higashi-cho 929, Kamogawa City, Chiba 296-8602, Japan.

**Keywords:** typical atrial flutter, entrainment, right atrial appendage, electrocardiogram, mapping

## Abstract

**Background:** Typical atrial flutter (AFL) is a macroreentrant tachycardia in which intra-cardiac conduction rotates counterclockwise around the tricuspid annulus. Typical AFL has specific electrocardiographic characteristics of a negative sawtooth-like wave in the inferior lead and a positive F wave in lead V1.

**Objective:** This study analyzed the source of the positive F wave in lead V1, which has not been well elucidated.

**Methods:** This study included 10 patients who underwent radiofrequency catheter ablation for a typical AFL. Electroanatomical mapping was performed both during typical AFL and entrainment from the right atrial appendage (RAA). The 12-lead electrocardiogram and three-dimensional (3D) electroanatomical maps were analyzed.

**Results:** The positive F wave in lead V1 changed during entrainment from the RAA in all the cases. The 3D map during entrainment from the RAA showed an area of antidromic capture around the RAA, which collided with the orthodromic wave in the anterior right atrium. This area of antidromic capture around the RAA was the only difference from the 3D electroanatomical map of tachycardia and is thought to be the cause of the disappearance of the F wave in lead V1 during entrainment.

**Conclusion:** The analysis of the differences in the 12-lead ECG and 3D maps between tachycardia and entrainment from the RAA clearly showed that activation around the RAA is responsible for the generation of the positive F wave in lead V1 of typical AFL.

## Introduction

Typical atrial flutter (AFL) is a commonly observed atrial arrhythmia with counterclockwise reentry of the tricuspid annulus^1^. It forms a circuit with the tricuspid annulus as an anatomical obstacle and terminal crest as a functional obstacle. A surface 12-lead electrocardiogram (ECG) is characterized by an asymmetric negative sawtooth-like wave in the inferior lead, positive F wave in lead V1, and negative F wave in lead V6.

Left atrial (LA) conduction is mainly involved in the sawtooth-like wave^2,3^. The waveform in lead V1 has been reported to reflect right atrial (RA) conduction by the analysis of the activation map in the three-dimensional (3D) mapping system^1^. In addition, in cavo-tricuspid isthmus (CTI)-dependent reentry, it has been reported that the polarity of the V1 lead changes when the direction of rotation changes^4,5^. These findings suggest that the positive F-wave in the V1 lead reflects conduction in the anterolateral RA wall. However, the correlation between surface ECG findings and intra-cardiac conduction has not been sufficiently investigated.

Entrainment provides useful information for the diagnosis and circuit localization of reentrant arrhythmias^6^. To date, there have been no reports on the visualization of intra-cardiac conduction during entrainment with a 3D mapping system. One report showed orthodromic and antidromic captured regions with intra-cardiac electrodes during a typical AFL^7^. This has the advantage of understanding temporal changes in intra-atrial conduction. However, it is nearly impossible to study the entire atrium simultaneously with the placed electrodes, and 3D construction is impossible. It may be difficult to understand whether the wavefront at the site of interest is orthodromic or antidromic during entrainment. Visualization using 3D mapping can thus deepen our understanding of entrainment.

In this study, we performed 3D mapping during entrainment in a typical case of AFL and investigated the source of the surface ECG waveform.

## Methods

This was a prospective, single-center observational study. This study was conducted in accordance with the principles of the Declaration of Helsinki. The study protocol was approved by our institutional review board (approval number, 22-021). Written informed consent was obtained from all patients.

### Patient population and data collection

Between April 2022 and May 2023, 10 patients with AFL or atrial fibrillation (AF) ablation who underwent mapping and ablation of typical AFL at Kameda Medical Center were included in this study. Patients between 20 and 80 years old with symptomatic AF or AFL refractory to antiarrhythmic drugs were eligible for inclusion in this study. The exclusion criteria were severe left ventricular systolic dysfunction (ejection fraction <35%), LA diameter >55 mm, prior cardiac surgery, and pregnancy. Patient information, including the medical history, echocardiographic findings, blood test results, procedure characteristics, and adverse events after the procedure, was collected from medical records. On transthoracic echocardiography, the LA diameter was defined as the maximum LA short-axis diameter in the parasternal view, and the LA volume was calculated from the apical view using the modified Simpson method.

### Mapping and catheter ablation procedure

All procedures were performed under general anesthesia using intubation for airway management, mechanical ventilation, intravenous and gas anesthesia, and muscle relaxation. If the patients were taking a direct oral anticoagulant (DOAC) other than dabigatran, the DOAC was switched to dabigatran after hospitalization. Warfarin or dabigatran was uninterrupted before and after the procedure. A decapolar catheter (CS EZ steer catheter, Biosense Webster, Diamond Bar, CA, USA; EPstar Snake, Japan Lifeline Co.,Ltd., Tokyo, Japan; Luma Cath, St. Jude Medical, Zaventem, Belgium) or duo-decapolar catheter (BeeAT; Japan Lifeline) was used to cannulate the coronary sinus. In the case of atrial fibrillation, a single transseptal puncture was performed under intra-cardiac echocardiography guidance. Prior to transseptal puncture, intravenous unfractionated heparin was administered to maintain an activated clotting time >300 s. A 3D navigation system (CARTO Mapping System; Biosense Webster) and a 20-electrode mapping catheter (PENTARAY NAV eco Catheter; Biosense Webster) or 64-electrode mapping catheter (OCTARAY Mapping Catheter; Biosense Webster) were used to create an electro-atomical map of the RA and/or LA. A 3D map was created during typical AFL and tachycardia entrainment from the right atrial appendage (RAA) using an ablation catheter (THERMOCOOL SMARTTOUCH SF Catheter; Biosense Webster) with an output of 5 to 10 V/2 ms. The surface 12-lead ECG was also recorded during both AFL and entrainment.

An open-irrigated ablation catheter (THERMOCOOL SMARTTOUCH SF Catheter; Biosense Webster) was used for ablation. In all cases, a deflectable sheath (Agilis NxT Steerable Sheath, St. Jude Medical, Zaventem, Belgium; SureFlex Steerable Guiding Sheath, Boston Scientific, Marlborough, MA, U.S.A; CARTO VizigoTM Bi-Directional Guiding Sheath; Biosense Webster) was used to achieve catheter stability. The CTI block line creation was performed using point-by-point ablation. The RF power was set at 35 W. RF ablation was applied in power-controlled mode with an irrigation rate of 2 ml/min during mapping and 30 ml/min during RF. A normal saline solution (NaCl 0.9%) was used. A bidirectional CTI block was confirmed in all cases using previously reported methods^8^.

### Analyses of 3D maps and surface 12-Lead ECGs

Activation maps during typical AFL showing intra-cardiac conduction were obtained with simultaneous recording of 12-lead ECGs. The positive F-wave in lead V1 was divided into ascending, peak, and descending segments and compared with intra-cardiac conduction at each time point. In addition, the sawtooth-like wave in the inferior lead was divided into slow descending, acute descending, and acute ascending portions, and the intra-cardiac conduction at each time point was compared. The areas of antidromic and orthodromic conduction were analyzed on 3D maps during entrainment from the RAA. The sites where antidromic and orthodromic conduction collided were visualized. The ECG during entrainment from the RAA and that during AFL were compared, and the differences in intra-cardiac conduction were analyzed.

### Statistical analyses

Descriptive statistics are reported as the mean ± standard deviation (SD) for continuous variables with a normal distribution, medians, and 25^th^ to 75^th^ percentiles for continuous variables with a skewed distribution. Categorical data were expressed as absolute values and percentages. The normality of distribution was tested using the nonparametric Kolmogorov-Smirnov test. Differences between the mean data were compared using a *t*-test for Gaussian variables, and the F-test was used to check the hypothesis of equality of variance. The Mann-Whitney nonparametric test was used to compare the non-Gaussian variables. Nominal variables were compared using the chi-square test or Fisher’s exact test, as appropriate. Statistical significance was set at P <0.05.

All statistical analyses were performed using the R software program, version 4.1.2 (EZR on R commander version 1.55).

## Results

### Study population and procedural parameters

Ten patients with AF or AFL underwent CTI ablations. Baseline demographic and procedural data of the 10 patients are presented in Table 1. In all cases, a complete bidirectional block line of CTI was achieved. No serious complications, including steam pop, cardiac effusion/tamponade, stroke, or esophageal injury, were observed.

**Table 1.** Baseline characteristics.

| **parameter** | **n=10** |
| --- | --- |
| Age (years) | 71±9 |
| Sex male, n (%) | 10 (100) |
| Body mass index (kg/m<sup>2</sup>) | 26.0±2.9 |
| Indication for ablation |  |
| Typical AFL, n (%) | 5 (50) |
| Paroxysmal AF, n (%) | 2 (20) |
| Persistent AF, n (%) | 3 (30) |
| LVEF (%) | 53±20 |
| LAD (mm) | 44.8±8.5 |
| LAV (ml) | 95.6±40.3 |
| Cardiomyopathy | 0 (0) |
| Hypertension, n (%) | 6 (60) |
| Diabetes mellitus, n (%) | 4 (40) |
| Prior stroke/TIA, n (%) | 1 (10) |
| Vascular disease, n (%) | 4 (40) |
| History of heart failure, n (%) | 3 (30) |
| CHADS<sub>2</sub> score | 1.6±1.4 |
| CHA<sub>2</sub>DS<sub>2</sub>-VASc score | 2.8±1.8 |
| Procedure duration (min) | 131.2±56.8 |
| Fluoroscopy time (min) | 11.3±8.7 |
| Complications during the procedure, n (%) | 0 (0) |
| Access site related, n (%) | 0 (0) |
| Cardiac tamponade, n (%) | 0 (0) |
| TIA/Stroke, n (%) | 0 (0) |
| Phrenic nerve palsy, n (%) | 0 (0) |
| Atrio-esophageal fistula, n (%) | 0 (0) |
| Death, n (%) | 0 (0) |

Values represent the mean ± SD or n (%).
AF, atrial fibrillation; AFL, atrial flutter; LVEF, left ventricular ejection fraction; LAD, left atrial diameter; LAV, left atrial volume; n, number; SD, standard deviation; TIA, transient ischemic attack

### Procedural details

RA mapping was performed during typical AFL in all cases, which revealed counterclockwise rotation around the tricuspid annulus (Figure 1 and 2). The median tachycardia cycle length (CL) of AFL was 234.2±29.7 ms. The negative sawtooth-like wave in the inferior leads and the positive F wave in the V1 lead were observed on ECGs in all cases. During AFL, entrainment from the RAA was performed. The pacing CL was 218.0±29.4 ms, which was 16.2±6.7 ms shorter than the AFL CL. RA mapping was performed during entrainment of the RAA. LA mapping was performed following RA mapping in 3 (30%) of the 10 cases (Figure 3).

**Figure 1.**
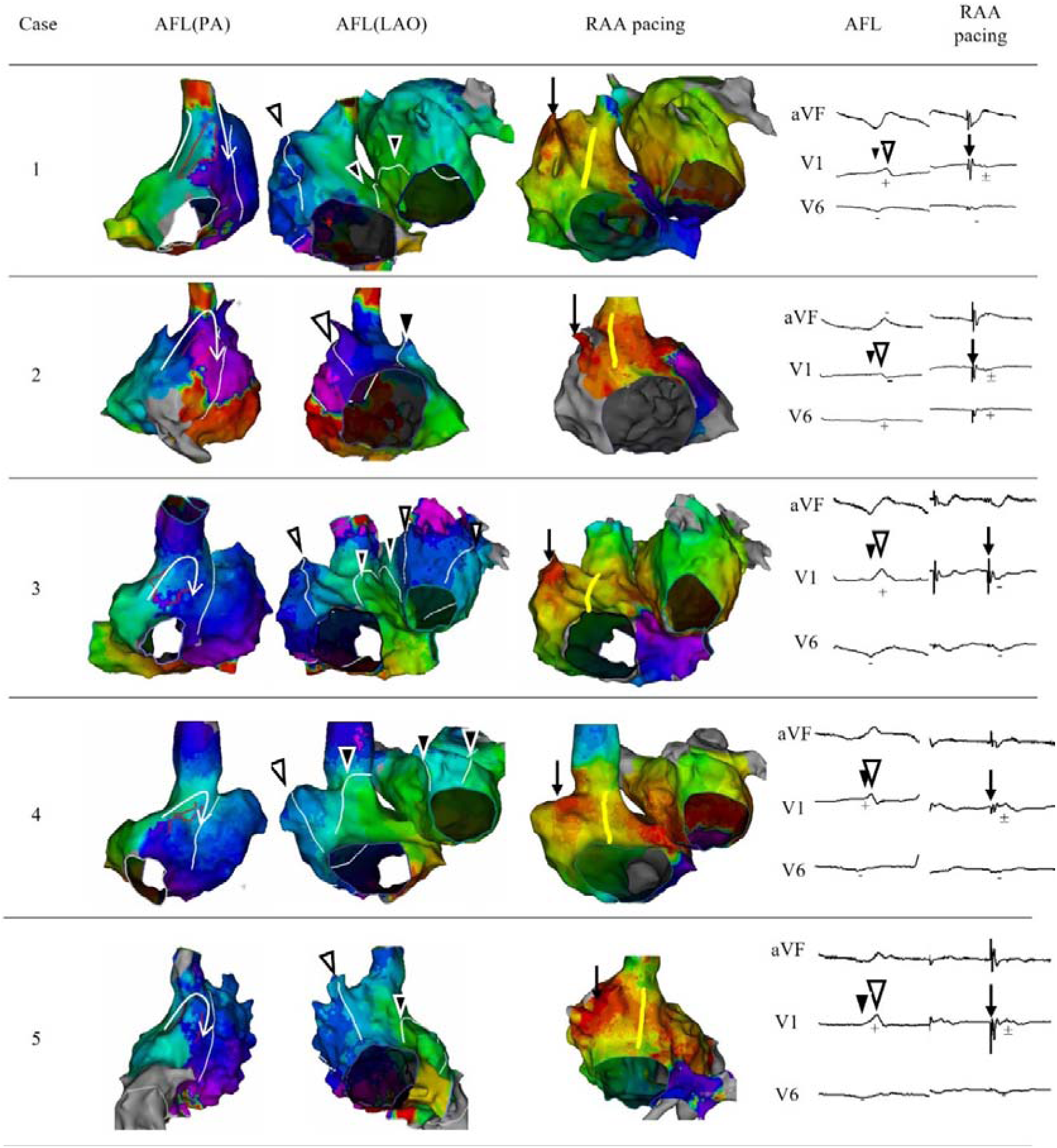
The activation map and ECG during both AFL and RAA pacing (cases 1-5) The activation map during AFL was compared with that during entrainment from the RAA. The white arrowhead in lead V1 represents the positive F peak. The white arrowhead on the 3D map represents the wavefront of the AFL conduction at the peak of the positive F wave. The black arrowhead in the ECG represents the beginning of the positive F-wave in lead V1. The black arrowhead on the 3D map represents the wavefront of the conduction at the beginning of the positive F wave. The arrow in the ECG represents the pacing spike and indicates the location of pacing insertion on the 3D map. The yellow line on the 3D map during RAA pacing represents the location of the collision between the antidromic conduction and orthodromic conduction from the previous pacing. In the 12-lead ECG, the F wave in lead V1 was altered during RAA pacing in all cases. In the PA view, the white line represents the ridge of the terminal crest, and double potentials are observed in the area surrounded by the red line. In the posterior RA, all cases showed caudal to cranial conduction in the septal side of the double potentials and cranial to caudal conduction in the lateral side of the double potentials. The site where double potentials were observed was posterior to the ridge of the terminal crest. AFL, atrial flutter; RAA, right atrial appendage; ECG, electrocardiogram; RA, right atrium.

**Figure 2.**
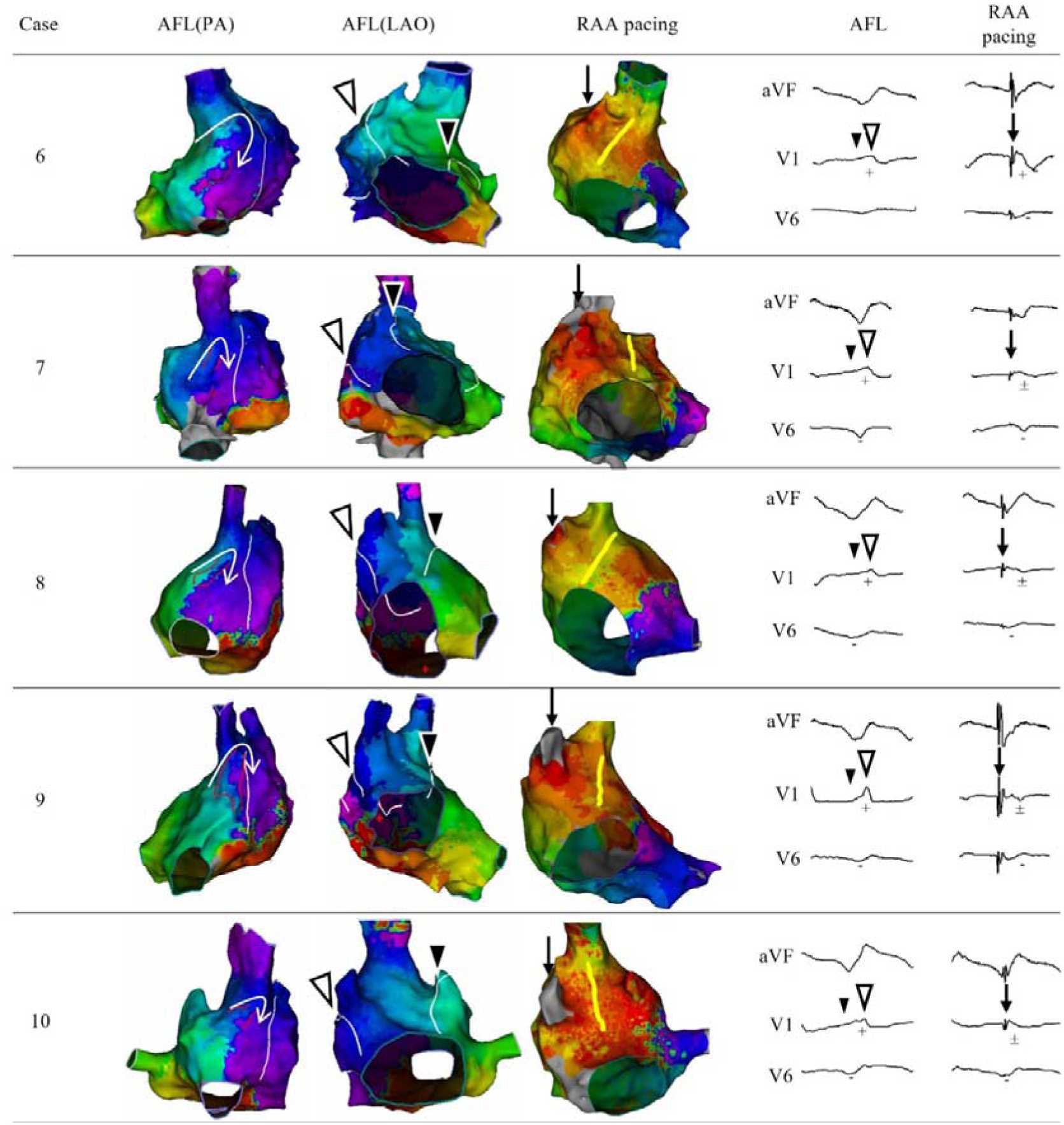
The activation map and ECG during both AFL and RAA pacing (cases 6-10)

**Figure 3.**
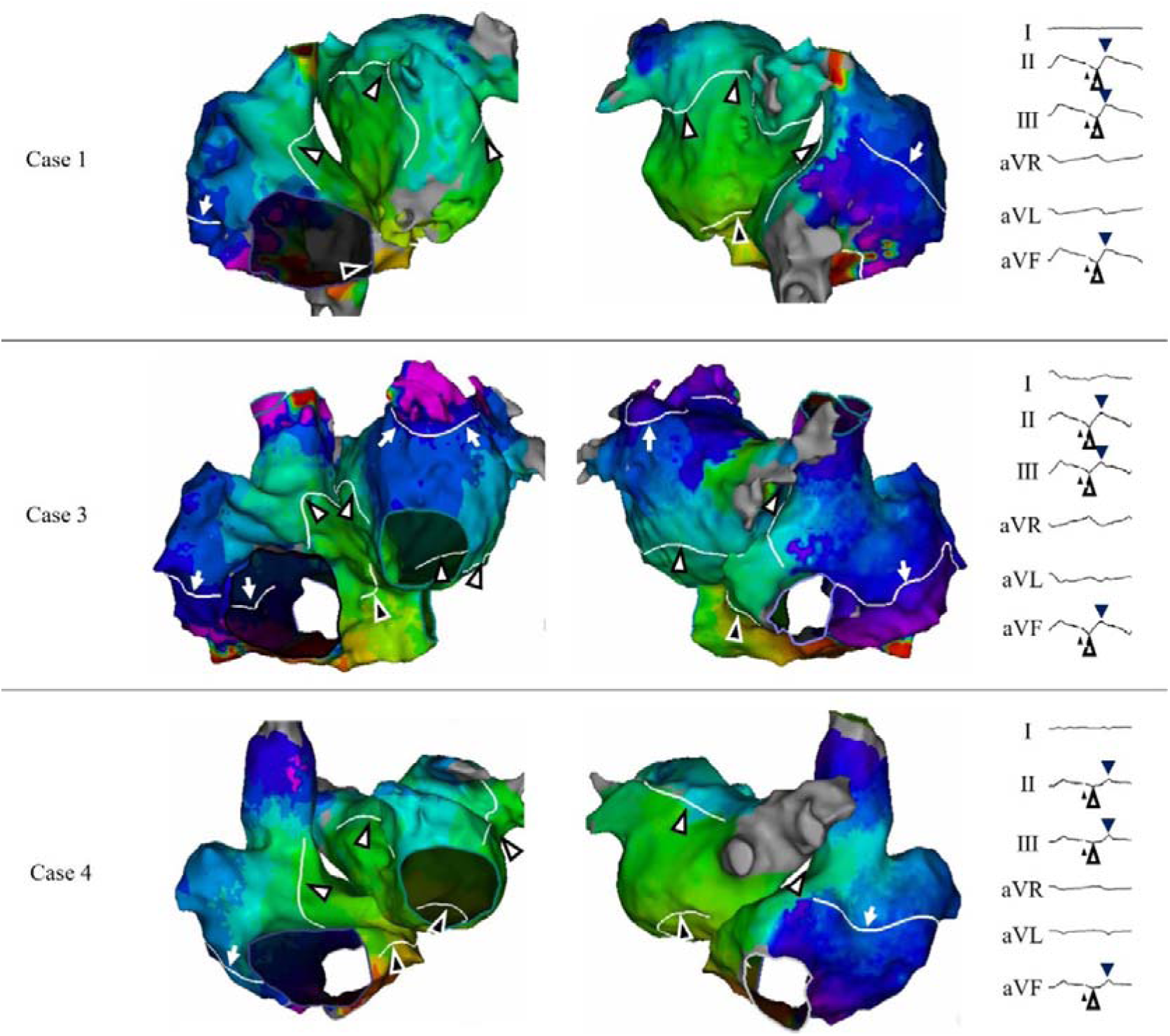
3D map of the RA and LA during AFL. Three-dimensional mapping of both the RA and LA during AFL was performed in 3 of the 10 cases. The beginning of the acute descending portion of the sawtooth-like wave in the inferior lead observed during AFL is indicated by a black arrowhead, and the endpoint is indicated by a white arrowhead. The black arrowheads on the 3D map represent the wavefront corresponding to the beginning of the acute descending portion of the sawtooth-like wave, and the white arrowheads represent the wavefront corresponding to the end of the acute descending portion of the sawtooth-like wave. AFL, atrial flutter; RA, right atrium; LA, left atrium.

### 3D mapping during AFL

A 3D map of the RA was acquired during AFL in all patients. The mean number of acquired points was 21878±11393. A 3D map of the LA was also acquired in three cases. The mean number of points acquired for the 3D map of LA was 12345±5777. In all cases, the local activation time (LAT) was continuously monitored, showing counterclockwise rotation around the tricuspid annulus and filling the total tachycardia cycle in the right atrium. The tricuspid annulus served as the anatomic obstacle, and the terminal crest served as the functional obstacle (Figures 1, 2, and 4). LAT of the LA filled only 64.1%±13.0% of the tachycardia cycle, with a passively activated pattern, with earliest activation located at the inferior LA septum (Figures 3 and 4).

**Figure 4.**
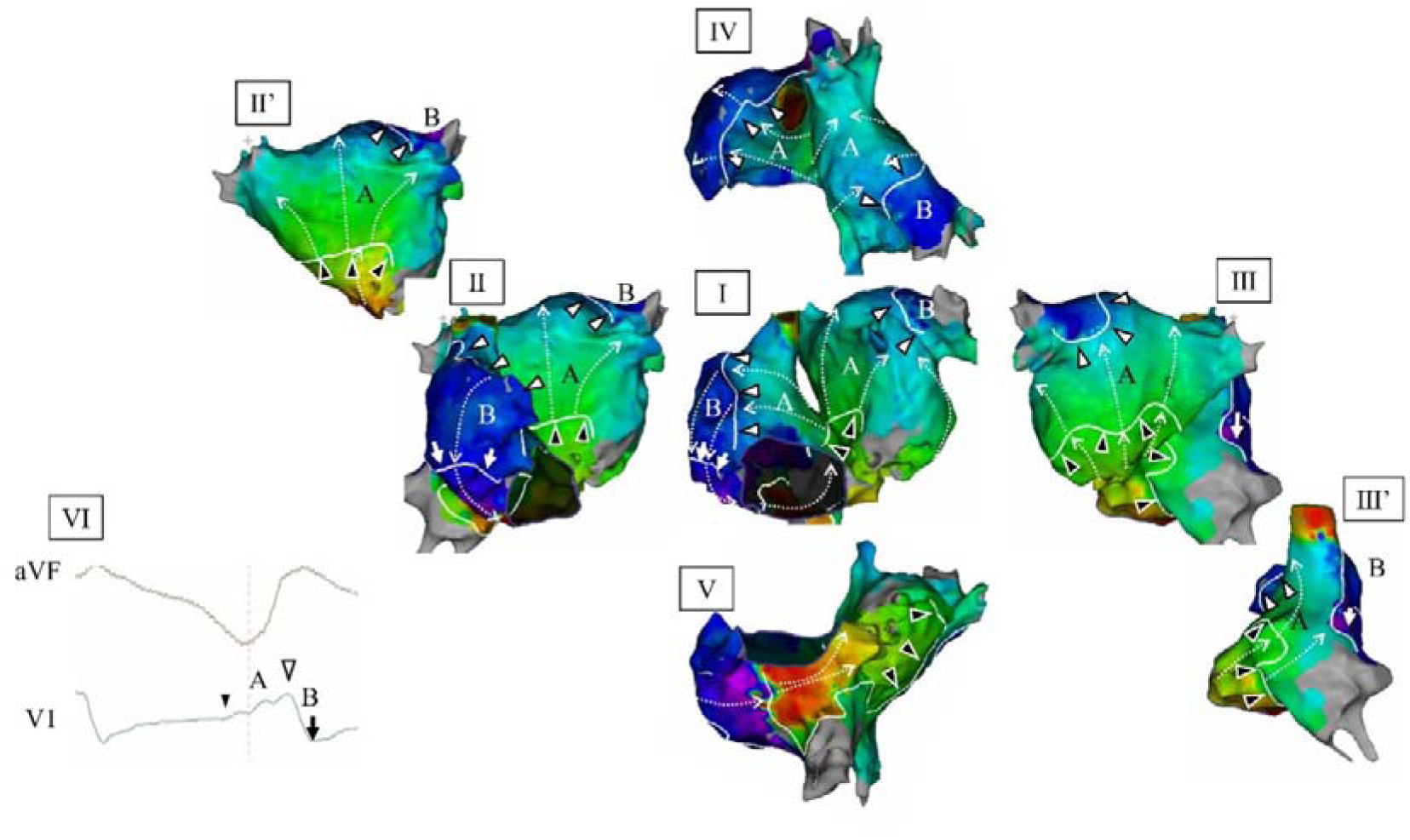
A representative case (case 1) The relationship between the activation map during AFL and ECG morphology of lead V1 is shown in this figure. Counterclockwise rotational conduction around the tricuspid annulus and terminal crest was observed in the RA. In the LA, caudal to cranial conduction was observed in the LA septum, lateral wall of LA, and posterior wall of LA. Conduction in the LA collided with the LA roof and LA appendage. The white arrowhead represents the peak of the positive F-wave in the V1 lead. The beginning of the F wave is indicated by a black arrowhead, and the endpoint is indicated by an arrow. The peak of the F-wave of lead V1 correlated with the ridge of the RAA. AFL, atrial flutter; RAA, right atrial appendage; ECG, electrocardiogram; RA, right atrium; LA, left atrium.

In the posterior RA, all cases showed caudal to cranial conduction in the septal side of the double potentials and cranial to caudal conduction in the lateral side of the double potentials. The site where double potentials were observed was posterior to the ridge of the terminal crest (Figures 1 and 2).

The slow descending portion of the sawtooth-like wave in the inferior lead was consistent with lateral-to-septal conduction of the RA inferior wall with no conduction in the LA. The acute descending portion of the sawtooth-like wave was consistent with the ascending conduction from the RA septum to the anterior wall. In the LA, this was consistent with ascending conduction in the septal, lateral, and posterior walls (Figure 3). The acute ascending portion of the sawtooth wave in the inferior lead was consistent with craniocaudal conduction across the anterolateral and posterolateral RA through the RAA in the RA and conduction to the collision site in the LA roof or LAA in the LA.

The peak of the positive F-wave in lead V1 corresponded to the activation timing at the superior ridge of the RAA (Figures 1, 2, and 4). The ascending portion before the peak of the positive F waves in the V1 lead corresponded to conduction along the anterior RA toward the superior ridge of the RAA, and the descending portion coincided with conduction along the lateral RA from the superior ridge of the RAA toward the inferior RA. In the LA, the ascending part of the positive F wave in the V1 lead corresponded with the conduction converging on the LA roof and appendage, which collided and disappeared before the activation wavefront reached the ridge of the RAA. There was no excitation in the descending portion of the left atrium (Figure 3).

Because the timing of the disappearance of LA conduction coincided with conduction from the RAA to the RA lateral wall, the peak of the positive F wave of the V1 lead almost corresponded to the peak of the positive overshoot component in the inferior leads (9/10 cases).

### 3D mapping during RAA entrainment

During typical AFL, constant pacing was delivered from the RAA at a pacing cycle 16.2±6.7 ms shorter than the AFL CL to entrain the tachycardia (Figures 1 and 2). Pacing was stopped as needed to confirm that the tachycardia persisted. The entrainment was confirmed with the findings of constant fusion in 3D mapping, as well as the F wave in 12-lead ECG. The post-pacing interval (PPI) was 279.6±27.7ms (AFL CL-PPI 45.4±13.4 ms). 3D mapping was performed during entrainment from the RAA during AFL. The RA was mapped in all cases. The mean number of points acquired was 16726±7707.

The paced wavefront fused with tachycardia and formed orthodromic conduction from the ridge of the RAA down the lateral RA toward the inferior RA, crossed the CTI, and reached the RA septum (Figures 1, 2, and 5). Conduction after crossing the CTI was identical to that during AFL and was considered orthodromic capture.

**Figure 5.**
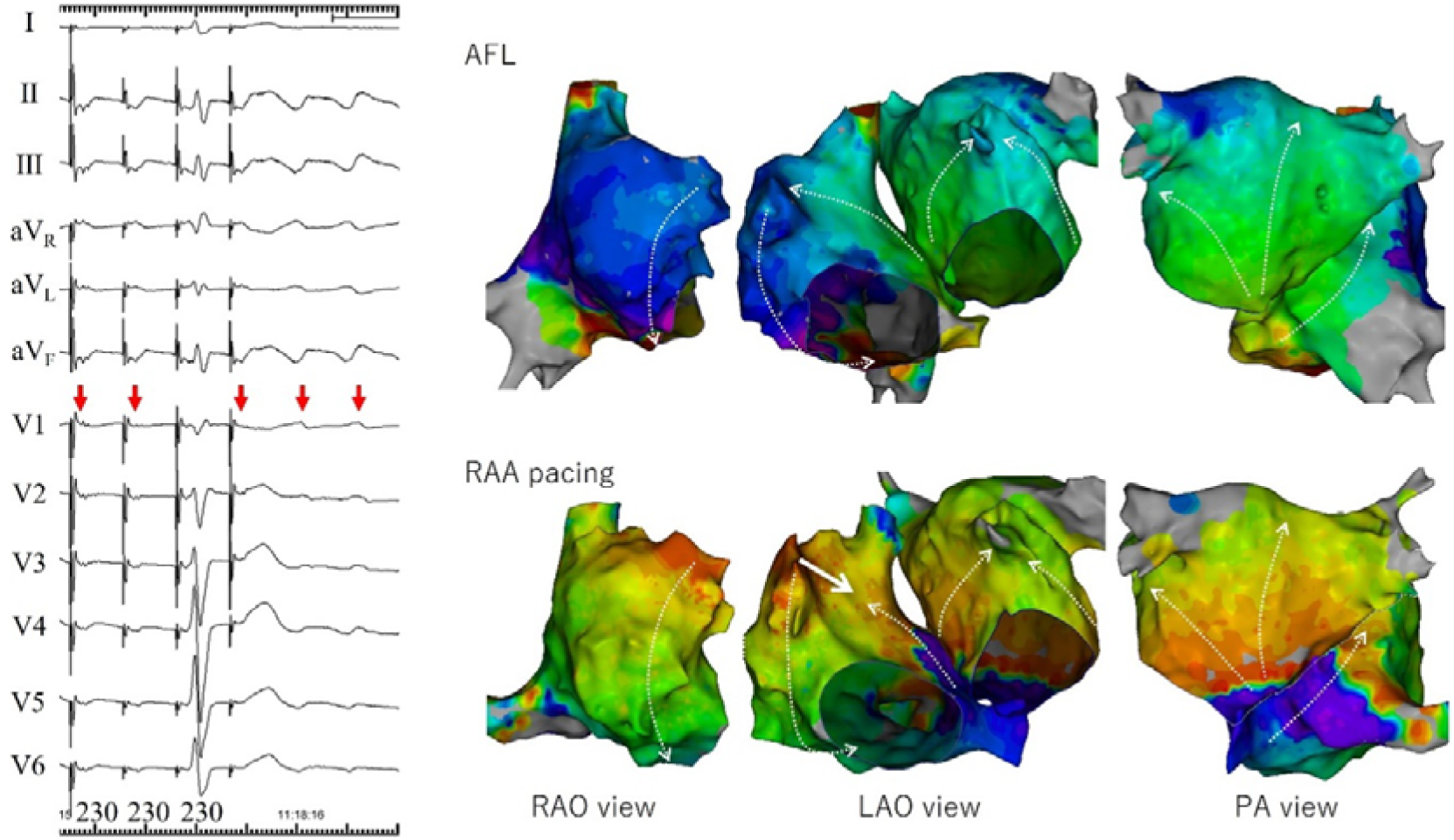
The comparison of AFL and pacing from the RAA in case 1. During entrainment from the RAA, the conduction in the lateral RA went cranial to caudal, passing through the cavo-tricuspid isthmus in the inferior RA, and then travelling cranially in the septum. This orthodromic conduction collided with the subsequent antidromic conduction in the anterior RA. The conduction pattern in the LA did not change compared with that during AFL. The positive F wave in lead V1 disappeared during RAA pacing. AFL, atrial flutter; RAA, right atrial appendage; RA, right atrium; LA, left atrium; RAO, right anterior oblique; LAO, left anterior oblique; PA, posterior-anterior.

In contrast, pacing created antidromic conduction from the RAA to the anterior RA, which collided with the orthodromic wavefront at the anteroseptal RA. The shortest time for antidromic conduction to collide with orthodromic conduction was 35.4±8.9 ms. LA 3D mappings were obtained in three cases. The mean number of points acquired was 12345±5777. In the LA, the conduction was identical to that during AFL without any component of antidromic capture, with the earliest activation occurring at the inferior septum and convergence at the LA roof and appendage (Figures 1, 2, and 5).

### ECG morphology and intra-cardiac conduction

A positive F wave was observed in lead V1 during AFL. There were eight cases of monophasic positive waves and two cases of biphasic positive/negative waves. The peak of the positive F-wave corresponded to the anterior ridge of the RAA in all cases. The local activation time difference between the peak of the positive F wave and the superior ridge of the RAA was only 4.0±3.9 ms. Furthermore, during entrainment from the RAA, disappearance of the positive F wave of lead V1 was observed in all cases (Figures 1, 2, and 5). There were nine cases of negative F waves in lead V6 during the AFL. During entrainment from RAA, the morphology of the negative F wave in lead V6 did not change.

The sawtooth-like wave in the inferior lead during AFL can be divided into gradually descending, acute descending, and acute ascending portions. The acute descending segment corresponds to ascending conduction of the RA septum. However, the corresponding areas varied from case to case. LA mapping was performed in only three cases. The acute descending segment corresponds to the ascending conduction in the LA septum, posterior wall, and lateral wall. In all cases, pacing from the RAA did not alter the acute descending segment.

### ECG morphologic change and intra-cardiac conduction during entrainment from the RAA

On comparing the 3D maps during typical AFL and entrainment from the RAA, the direction of the wavefront changed only in the area of antidromic conduction in the anterior RAA. The area of orthodromic conduction was nearly unchanged with intra-cardiac conduction during the AFL. The positive F-wave in lead V1 disappeared during entrainment from the RAA. Since intra-cardiac conduction is the cause of the ECG morphology, we concluded that, in typical AFL, positive F waves in lead V1 are caused by conduction around the RAA.

## Discussion

### Major findings

In the present study, we established the 3D map during entrainment of the AFL from the RAA and the relationship between intra-cardiac conduction and the surface ECG in typical AFL. The results of this study are as follows: (1) The 3D map during entrainment clearly shows antidromic and orthodromic conduction, which are constantly fused. (2) The positive F wave in lead V1 during a typical AFL corresponds to conduction around the RAA. (3) RAA entrainment alters the morphology of the F wave in lead V1 without major changes in the inferior and V6 leads.

### 3D mapping of AFL

Shah et al. created a 3D map of typical AFL and detailed intra-atrial conduction^9^. They visualized the tricuspid annulus and terminal crest as re-entry obstacles. However, the correlation between the ECG and intra-cardiac conduction has not been clarified. In recent years, simultaneous multipoint mapping and high-density mapping have become possible, and the accuracy of 3D mapping has consequently improved. However, there have been no reports of 3D mapping during entrainment of reentrant tachycardia.

It is possible to clearly demonstrate orthodromic and antidromic conduction, which is useful for understanding reentry. Our study showed the entire AFL circuit in 10 patients. A reentrant circuit existed around the tricuspid annulus. However, the ridge of the terminal crest is not thought to be a functional obstacle in AFL. If we interpret the site of double potential as the site of the functional block, the border between the terminal crest and sinus venarum seems to serve as a functional obstacle.

### Generation of F wave in the lead V1

An examination of the correlation between 3D mapping during AFL and 12-lead ECGs showed that the peak of the positive F wave in lead V1 was precisely correlated with the timing of conduction through the ridge of the RAA. By performing entrainment from the RAA during AFL, the 3D map showed antidromic conduction from the RAA to the septal side, and orthodromic conduction from the RAA to the lateral RA, CTI, septum, and anterior RA. Compared to the 3D map during AFL, the area of antidromic conduction was from the ridge of the RAA to the anterior RA. During entrainment from the RAA, the positive F waves in lead V1 changed in all the cases. The difference in the direction of conduction during AFL and entrainment from the RAA was only in the area of antidromic conduction, which was interpreted as the cause of changes in the ECG.

These findings are consistent with those of previous studies. Notaristefano et al. reported that the morphology of lead V1 and conduction in the lateral RA were consistent in 3D map^1^, and our present study had similar results. In addition, Saoudi et al. reported that differences in the direction of rotation led to changes in the polarity of F waves in CTI-dependent reentry^5^. However, in the present study, we demonstrated that the direction of conduction around the RAA had an even greater contribution, and we limited the areas affecting the waveform in lead V1.

Focal atrial tachycardia (AT) originating from the LA reportedly has a positive P’ wave in lead V1^10,11^. Conduction from the LA to the RA often passes through Bachman’s bundle, going in the medial to lateral direction in the RAA and cranial to caudal direction in the lateral RA. Therefore, it is considered consistent that the polarity of lead V1 is positive, even in AT originating from the LA. It has also been reported that the ECG of focal AT arising from the RAA shows a negative P’ in lead V1. Focal AT from the RAA shows conduction away from the RAA. The fact that the polarity of the morphology of lead V1 reflects conduction around the RAA in AT arising from the RAA is consistent with the present findings.

### The relationship between the sawtooth-like wave of the inferior lead and the conduction in the RA

Entrainment from the RAA did not allow the antidromic conduction zone to reach the RA septum. Therefore, conduction in the LA through Bachman’s bundle is not affected during RAA pacing. When the tachycardia cycle is shortened by entrainment, conduction in the RA septum is altered in some cases, but the morphology of the inferior leads is not altered. The characteristic morphology of a sawtooth-like wave is an acute descending portion. This morphology was consistent with conduction simultaneously ascending through the RA septum, LA septum, LA posterior wall, and LA lateral wall. In an observational study of AFL in dogs and an examination of the morphology of the inferior lead, Okumura et al. showed that the LA contributed to the ECG morphology of AFL^3^. The results of the present study are consistent with those of that previous report.

### Intra-cardiac conduction during entrainment

When entrainment is performed from the RAA during AFL, and a 3D map is created during entrainment, it is possible to clearly identify the areas of antidromic and orthodromic capture. During entrainment, AFL conduction and conduction due to RAA pacing fused in the anterior and lateral RA. Because the CTI forms an anatomical isthmus, conduction passing through the CTI is almost identical to that of the AFL. The area of activated conduction through the CTI is defined as the orthodromically captured region. In this case, a conduction delay is not required to define the orthodromically captured region. In addition, although the conduction of the lateral RA is a fusion of the pacing and AFL wavefronts, the conduction direction can be interpreted as orthodromic. When observing the direction of intra-cardiac conduction using intra-cardiac electrodes^12^, interpreting whether an area is fused or orthodormically captured can be difficult. The 3D map allows the observation of intra-cardiac conduction during entrainment and may be useful for understanding entrainment.

### Limitations

Several limitations associated with the present study warrant mention. First, it was conducted at a single center and included a small sample size. However, the purpose of this study was to clarify intra-cardiac conduction during entrainment and to evaluate ECG changes during entrainment; the results were consistent in all cases, so it may be possible to draw conclusions even with a small number of cases. Second, we included patients with atrial fibrillation and AFL. The degree of atrial remodeling was not uniform, and some cases may have been atypical in terms of ECG changes. Third, only three patients underwent LA mapping, suggesting that our findings may lack generalizability regarding LA involvement.

## Conclusion

By creating a 3D map during entrainment of a typical AFL, it was possible to clearly observe the orthodromically captured region and the antidromically captured region. It was found that conduction around the RAA had a major effect on the morphology of the V1 lead as depicted by the 12-lead ECG. In addition, it was suggested that conduction in the LA played a major role in the generation of sawtooth-like waves in the inferior lead.

## Date availability

Date openly available in Zendo repository, reference number 8150179.

## Ethics approval

Kameda Medical Center institutional review board.

